# Dynamic MR of muscle contraction during electrical muscle stimulation as a potential diagnostic tool for neuromuscular disease

**DOI:** 10.1101/2024.09.17.24313673

**Authors:** Francesco Santini, Michele Giovanni Croce, Xeni Deligianni, Matteo Paoletti, Leonardo Barzaghi, Niels Bergsland, Arianna Faggioli, Giulia Manco, Chiara Bonizzoni, Ning Jin, Sabrina Ravaglia, Anna Pichiecchio

**Affiliations:** Basel Muscle MRI, Department of Biomedical Engineering, University of Basel, Basel, Switzerland; Department of Radiology, University Hospital Basel, Basel, Switzerland; Department of Brain and Behavioral Sciences, University of Pavia, Pavia, Italy; Advanced Imaging and Artificial Intelligence Research Unit, Neuroradiology Department, IRCCS Mondino Foundation, Pavia, Italy; Department of Neurology, Jacobs School of Medicine and Biomedical Sciences, Buffalo Neuroimaging Analysis Center, University of Buffalo, The State University of New York, Buffalo, New York, USA; Istituti Clinici Scientifici Maugeri IRCCS, Servizio di Diagnostica per Immagini - Istituto di Montescano, Italy; Cardiovascular MR R&D, Siemens Medical Solutions USA, Inc., Cleveland, OH, USA; IRCCS Mondino Foundation, Pavia, Italy

## Abstract

Thanks to the rapid evolution of therapeutic strategies for muscular and neuromuscular diseases, the identification of quantitative biomarkers for disease identification and monitoring has become crucial. Magnetic resonance imaging (MRI) has been playing an important role by noninvasively assessing structural and functional muscular changes. This exploratory study investigated the potential of dynamic MRI during neuromuscular electrical stimulation (NMES) to detect differences between healthy controls (HCs) and patients with metabolic and myotonic myopathies. The study included 14 HCs and 10 patients with confirmed muscular diseases. All individuals were scanned with 3T MRI with a protocol that included a multi-echo gradient echo sequence for fat fraction quantification, multi-echo spin-echo for water T2 relaxation time calculation, and 3D phase contrast sequences during NMES. The strain tensor, buildup and release rates were calculated from velocity datasets. Results showed that strain and strain buildup rate were reduced in the soleus muscle of patients compared to HCs, suggesting these parameters could serve as biomarkers of muscle dysfunction. Notably, there were no significant differences in fat fraction or water T2 measurements between patients and HCs, indicating that the observed changes reflect alterations in muscle contractile properties that are not reflected by structural changes. The findings provide preliminary evidence that dynamic muscle MRI during NMES can detect abnormalities in muscle contraction in patients with myotonia and metabolic myopathies, warranting further research with larger, more homogeneous patient cohorts.

## Introduction

Neuromuscular diseases (NMDs) usually affect children from a young age and can be fatal. The most common causes of neuromuscular diseases are rare genetic conditions that have long been untreatable. In recent years, however, new therapies (including gene therapy) are opening new doors for affected individuals, albeit for a limited number of known diseases (1–5). With the development of novel therapies, the need for reliable biomarkers that can provide objective insight into the course of the disease and detect early therapeutic effects has arisen.

In particular, magnetic resonance imaging (MRI) has been proven to provide exceptional flexibility and accuracy, with the capability of assessing muscle geometry and morphology, tissue composition, and function, over the whole organ of interest, in the same examination (6). However, despite the importance of MRI in characterizing NMDs, it is still unable to provide a truly comprehensive picture of the muscle composition and functionality, with the most critical aspect being the quantitative assessment of interstitial muscle fibrosis.

The evaluation of muscle status in dystrophic diseases has so far mostly relied on T1-weighted imaging for the assessment of chronic muscular changes and on fat-suppressed T2-weighted imaging for the assessment of the acute activity of the disease (7–9). While these methods are fast and able to provide a rough staging of the disease (10), they are increasingly complemented by quantitative assessments, that is, methods that can provide an objective quantification of the underlying tissue characteristics (11). Depending on the investigated pathology, two key techniques include fat-suppressed T2 quantification methods, given their ability to accurately describe the activity of the disease, being sensible to inflammatory edema (12,13), and three-dimensional fat/water quantification (14), which can potentially depict muscle morphology with great accuracy and can provide an objective measurement of how the muscle fibers have been permanently compromised.

While promising, however, these markers do not highlight all aspects of the pathology as they only focus on the inflammation and irreversible replacement stages (15). Accurate markers of other aspects of the muscular microstructure are needed to obtain a better understanding of the clinical course of various muscle pathologies and to achieve higher levels of accuracy in diagnosis and monitoring of disease progression.

Dynamic imaging of muscle contraction is one potential functional marker that has already been successfully applied in healthy individuals, especially in the context of aging (16–19). The derived parameters of strain and strain rate are sensitive to the age-related physiological changes of the muscle, resulting in a reduction of these two quantities. These parameters reflect the ability of the muscle to deform when performing its natural function of contracting to generate an output force.

In order to elicit a periodic contraction of the muscle, required for high-temporal-resolution imaging by using methods such as phase contrast acquisitions, neuromuscular electrical stimulation (NMES) can be used in synchronization with the acquisition, which can provide reproducible results (20,21). This particular technique has been recently used in a study on patients with facioscapulohumeral dystrophy (FSHD), yielding promising results on the possibility of using the derived parameters of strain and contraction and relaxation rates as functional biomarkers of disease (22). This study was however limited to a single slice acquisition of the thigh.

Due to preliminary evidence showing a reduction in strain and strain rate along with a decrease in functionality of the muscle, it can be hypothesized that these markers can also be reduced in neuromuscular diseases affecting muscle functionality. We analyzed muscular and neuromuscular diseases likely associated with changes in contraction parameters (stiffness, relaxation time), such as myotonias (dystrophic and non-dystrophic), as well as those in which repeated exercise is expected to cause transient weakness, such as metabolic myopathies. Thus, in this study, we are expanding the investigation to a wider spectrum of diseases affecting the skeletal muscle with an exploratory analysis of 10 patients with dystrophic and non-dystrophic myotonias and metabolic myopathies, and 14 healthy controls (HCs), focusing on time-resolved, three-dimensional acquisition of the triceps surae during electrical muscle stimulation.

## Materials and Methods

### Study population

Fourteen HCs (5 male, median age 50y, range: 35-62) and 10 patients (6 male, median age 46y range: 25-66) were prospectively recruited for this study. The patients had a confirmed diagnosis of muscular or neuromuscular disease, and specifically: myotonic dystrophy Type 2 and Type 1, chloride channel myotonia, SCN4A channelopathy, and McArdle disease. Among metabolic myopathies, we selected two patients with McArdle disease, since this disease is not associated with structural muscle degeneration but is rather characterized by transitory weakness induced by short-time exercise/muscle contraction. Among myotonias, we selected three patients with myotonic dystrophy in the early disease stage (without calf muscle wasting) and five with non-dystrophic myotonias caused by chloride channel mutations (3 patients) or sodium channel mutations (2 patients). Details on patients demographics, clinical features and diagnosis are in supplementary Table 1. Only one patient was taking medications for myotonia at the time of MRI. Since the MRI analysis was performed on lower limbs, the severity of myotonia as well as the functionality of lower limbs was assessed by the Timed-Up-and-Go test (TUG), in which the patients were instructed to stand up, walk 6 meters and sit back down again.

All subjects gave their written informed consent to participate. The study conformed to the Declaration of Helsinki, and the experimental procedures were approved by the local ethics committee.

### MR image acquisition and processing

During a preparation phase, two sets of gel-based electrodes were placed on the subject’s right leg. The stimulation intensity was tested, and the evoked force was measured before the scan with a custom-made MR-compatible sensor (23), as well as the maximum voluntary force (MVF) in the same position. The stimulation current was established immediately before the scan as the current required to elicit a force of approximately 10% of the MVF.

For the stimulation, a commercial 2-channel EMS device was used to induce periodic muscle contraction of the calf muscles. Biphasic stimulation with rectangular pulses was applied (pulse width: 400 μs, pulse frequency: 80 Hz, contraction duration: 750 ms, release duration: 750 ms release).

During the MR acquisition, the second channel of the stimulator was converted to a trigger signal by a custom electronic device (20).

The subjects were scanned on a 3T clinical MRI scanner (MAGNETOM Skyra, Siemens Healthineers, Forchheim, Germany). The scanning protocol included a multi-echo gradient-echo sequence for fat fraction quantification (axial 6-point Dixon gradient echo (GRE) sequence (matrix size = 432 × 396; TR = 35 ms; TE = 1.7–9.2 ms; resolution = 1.0 × 1.0 × 5mm3; scan time = 15 min), a multi-echo spin-echo sequence for the quantification of the T2 relaxation time of the water component (water T2 - TE/TR = [10.9-185.3] ms /4100.0 ms, 17 echo times; resolution = 1.2 × 1.2 × 10.0 mm3; slice gap = 30 mm; scan time = 5 min) (13), and, as last, a three-dimensional phase contrast sequence for the evaluation of the contraction during NMES.

For the dynamic acquisition, a prospectively-gated, highly accelerated cartesian 4D flow research sequence using L1-regularized wavelet-based compressed sensing (24,25) was placed in a sagittal orientation to cover the whole calf. The imaging protocol had the following parameters: TE/TR 4.1/8.7 ms, flip angle 10°, bandwidth 910 Hz/px, matrix size 128×54×48, resolution 2.3×2.3×2.5 mm^3^, Venc 15 cm/s, 2 k-space lines per segments, acceleration factor 7.6. The acquisition time was approximately 5 minutes per each dataset.

Fat fraction maps were calculated using the publicly-available FattyRiot algorithm (26).

Water T2 maps were calculated through a fitting of a dual-compartment signal model simulated with the extended-phase-graph method (12).

The velocity datasets were subsequently post-processed offline to calculate the strain tensor at each spatial location throughout the stimulation cycle (20). The largest positive eigenvalue of the strain tensor was considered for further quantitative processing, in line with (21,22). The “buildup” and “release” rates, related to the speed at which the maximum contraction was reached, or the speed of the return to the relaxed state, respectively, were calculated through a sigmoid fit of the eigenvalue time curve, as described in (19).

### Data analysis

The datasets were segmented using the computer-assisted segmentation software Dafne (27) and the average values over the soleus and the lateral and medial gastrocnemii were extracted of the following variables: first positive eigenvalue of the strain tensor, buildup rate, release rate, fat fraction, and water T2. The dynamic parameters were averaged over the volume of the muscle between the centers of the two NMES electrodes, as identifiable in the MR image.

Data analysis was performed in R (28). Due to the non-normality of the distributions, the results are presented as median values and 1st and 3rd quartiles for each considered value. For each variable, the results of a standardized logistic regression modeling with the binarized subject group (HC or patient, where the “patient” was assigned a value of 1) as dependent variable, and the normalized (mean-detrended and divided by the standard deviation) considered variable as independent variable are presented. The outcome of the modeling is shown in terms of odds ratios and their 95% confidence interval.

Association between the dynamic-derived quantities (strain, and buildup and release rates) and the quantitative parameters (fat fraction and water T2) is investigated through Spearman’s correlation.

The analysis script and the extracted tabular data used are available at (29).

## Results

Both HCs and patients had similar median levels of fat percentage in the three muscles between 3 and 4%, although three different patients had at least one muscle with a fat percentage above 10% (Figure 1a). Similarly, median water T2 values were between 34.3 and 36.0 for all muscle ROIs in both groups (Figure 1b). The median strain, however, was higher in HCs in all three muscles, with an increase of up to 36% (0.26 vs 0.19) in median value in the soleus of healthy volunteers with respect to patients, and 39% (but with higher variability) in the lateral gastrocnemius (Figure 2a). The buildup rate was slightly faster in the soleus (0.05 vs 0.03 s-1) but not in the other muscles (Figure 2b and 2c). The values are summarized in table 1.

**Table 1:** Values of the considered variables for the three muscles of interest for healthy controls and patients. The last column represents the odds ratios of the logistic regression model applied to the variable. NA represents a value that could not be computed by the model.

| **ROI** | **Variable** | **Healthy controls**<br>(Median and interquartile range) | **Patients**<br>(Median and interquartile range) | **Odds ratio**<br>(estimate and 95% confidence intervals) |
| --- | --- | --- | --- | --- |
| **Soleus** | **Fat fraction** | 0.04 (0.03 - 0.04) | 0.04 (0.03 - 0.05) | 1.83 (0.77 - 10.01) |
|  | **Water T2 [ms]** | 35.5 (35.0 - 36.2) | 35.0 (34.6 - 36.6) | 0.73 (0.32 - 1.54) |
|  | **Strain** | 0.26 (0.17 - 0.31) | 0.19 (0.12 - 0.21) | 0.34 (0.10 - 0.89) |
|  | **Buildup rate [s<sup>-1</sup>]** | 0.045 (0.041 - 0.061) | 0.031 (0.030 - 0.042) | 0.38 (0.11 - 0.99) |
|  | **Release rate [s<sup>-1</sup>]** | 0.013 (0.011 - 0.014) | 0.012 (0.010 - 0.013) | 0.88 (0.37 - 2.08) |
| **Medial Gastrocnemius** | **Fat fraction** | 0.03 (0.03 - 0.04) | 0.03 (0.03 - 0.05) | 4.96 (0.81 - NA) |
|  | **Water T2 [ms]** | 36.0 (34.7 - 36.4) | 34.3 (33.9 - 36.0) | 0.29 (0.05 - 0.96) |
|  | **Strain** | 0.22 (0.14 - 0.33) | 0.20 (0.13 - 0.22) | 0.54 (0.19 - 1.30) |
|  | **Buildup rate [s<sup>-1</sup>]** | 0.044 (0.030 - 0.078) | 0.036 (0.032 - 0.062) | 0.73 (0.27 - 1.69) |
|  | **Release rate [s<sup>-1</sup>]** | 0.010 (0.008 - 0.012) | 0.011 (0.009 - 0.012) | 1.81 (0.76 - 5.21) |
| **Lateral Gastrocnemius** | **Fat fraction** | 0.04 (0.03 - 0.04) | 0.03 (0.03 - 0.05) | 1.92 (0.70 - NA) |
|  | **Water T2 [ms]** | 36.0 (34.6 - 36.5) | 35.4 (33.9 - 36.4) | 0.93 (0.45 - 1.98) |
|  | **Strain** | 0.25 (0.16 - 0.31) | 0.18 (0.13 - 0.25) | 0.47 (0.17 - 1.13) |
|  | **Buildup rate [s<sup>-1</sup>]** | 0.034 (0.025 - 0.049) | 0.035 (0.030 - 0.040) | 0.76 (0.29 - 1.76) |
|  | **Release rate [s<sup>-1</sup>]** | 0.009 (0.008 - 0.014) | 0.009 (0.007 - 0.011) | 0.67 (0.24 - 1.56) |

**Figure 1:**
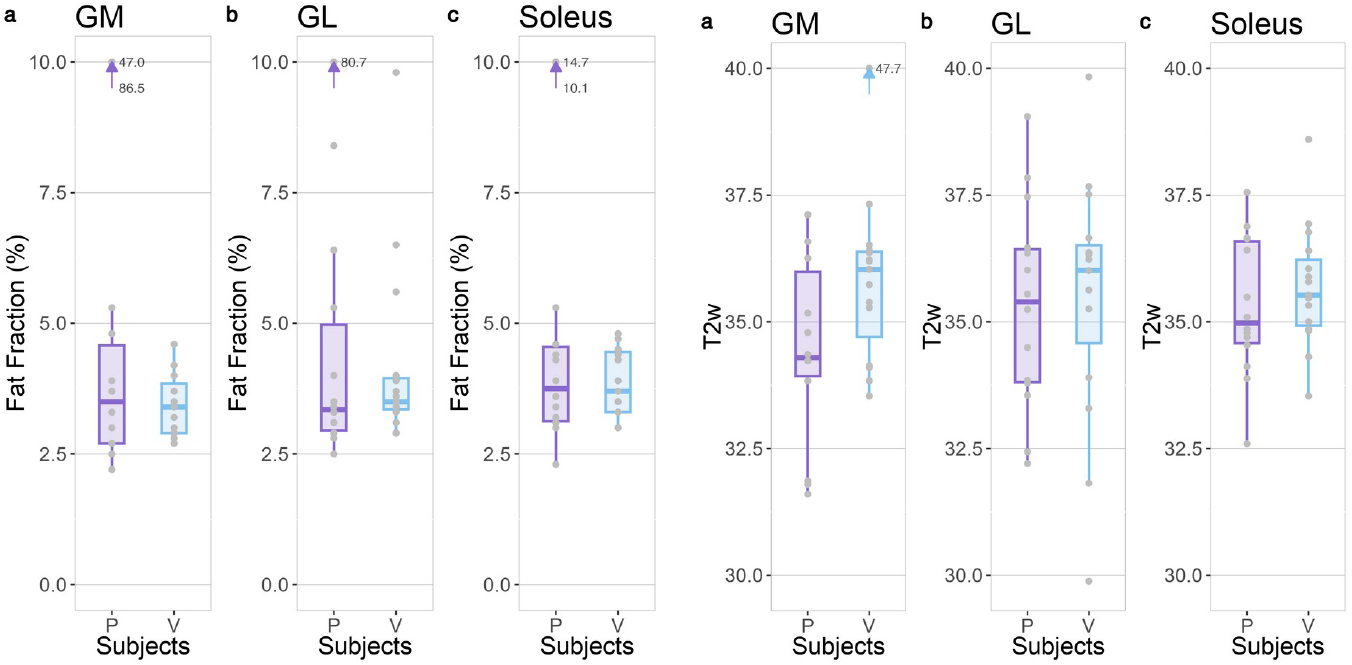
Fat fraction (left) and water T2 (right) values for each ROI in patients (P) and healthy volunteers (V). Outliers outside the limits of the plot are represented by an arrow and corresponding values.

**Figure 2:**
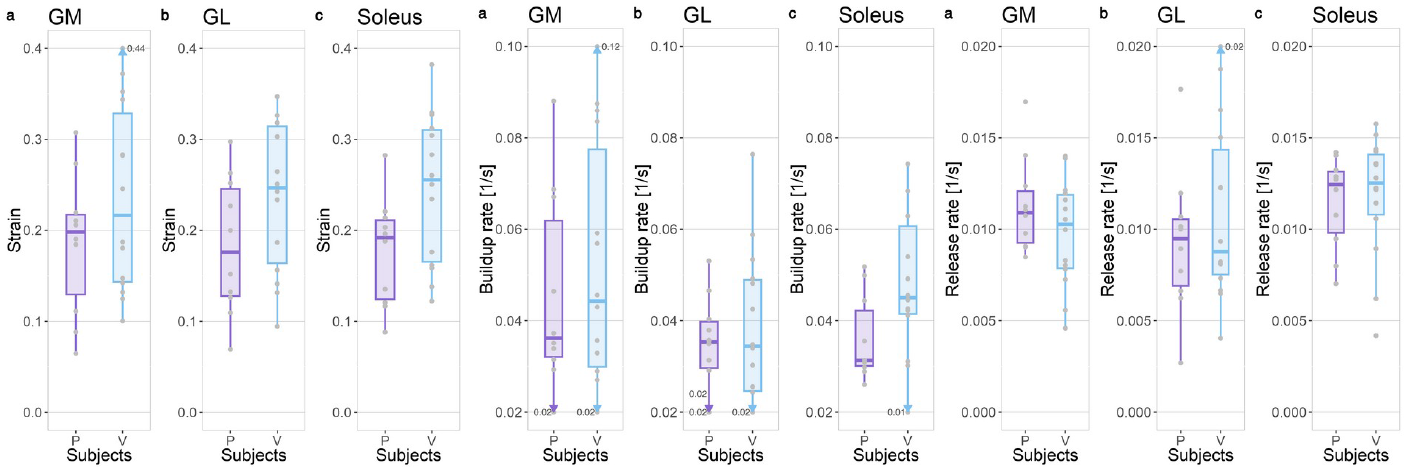
Strain (left), buildup rate (center), and release rate (right) for each ROI in patients (P) and healthy volunteers (V). Outliers outside the limits of the plot are represented by an arrow and corresponding values.

The logistic regression model showed a fitted odds ratio of 0.34 with 95% confidence interval of 0.10 to 0.89 for the strain value of the soleus, meaning that an increase of one standard deviation in the strain decreases the likelihood of a subject being in the “patients” group by a factor of 0.34. Similarly, the buildup rate in the soleus has a fitted odds ratio of 0.38 with 95% confidence interval 0.11 to 0.99. Although with lower confidence, and with a 95% confidence interval that crosses the identity line, increased fat fraction is also a compatible indicator of belonging to the patient group (Figure 3). The logistic regression for water T2 of the gastrocnemius medialis had an odd ratio of 0.29 (confidence interval 0.05 - 0.96), driven by a single outlier in the volunteer group with an non-physiologically high T2 value, possibly due to physical activity before the scan, or image artifact.

**Figure 3:**
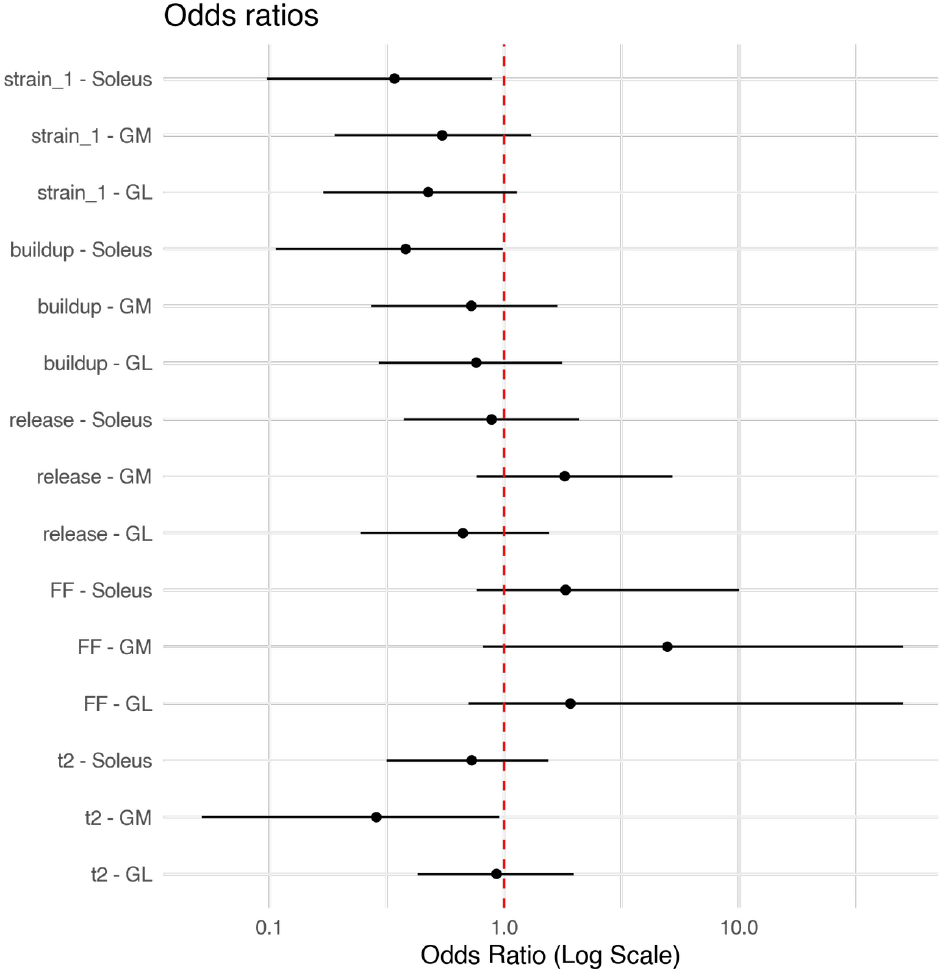
Odds ratios of the considered variables across the three ROIs with 95% confidence interval. An odds ratio of 1 means that an increase in the considered variable has no influence on the likelihood that the subject belongs to the “patient” or “healthy control” group. A higher odds ratio means that an increase in the variable increases the likelihood for the subject to be a patient, and vice versa for odds ratios lower than 1.

Correlations between the quantitative and dynamic variables were negligible, with the highest correlation observed being of −0.188 between the strain and fat fraction (table 2 and figure 4).

**Table 2:** Spearman’s correlation coefficients between the dynamic and quantitative variables.

|  | **Fat fraction** | **Water T2** |
| --- | --- | --- |
| **Strain** | -0.188 | -0.276 |
| **Buildup rate** | 0.103 | -0.060 |
| **Release rate** | 0.033 | -0.192 |

**Figure 4:**
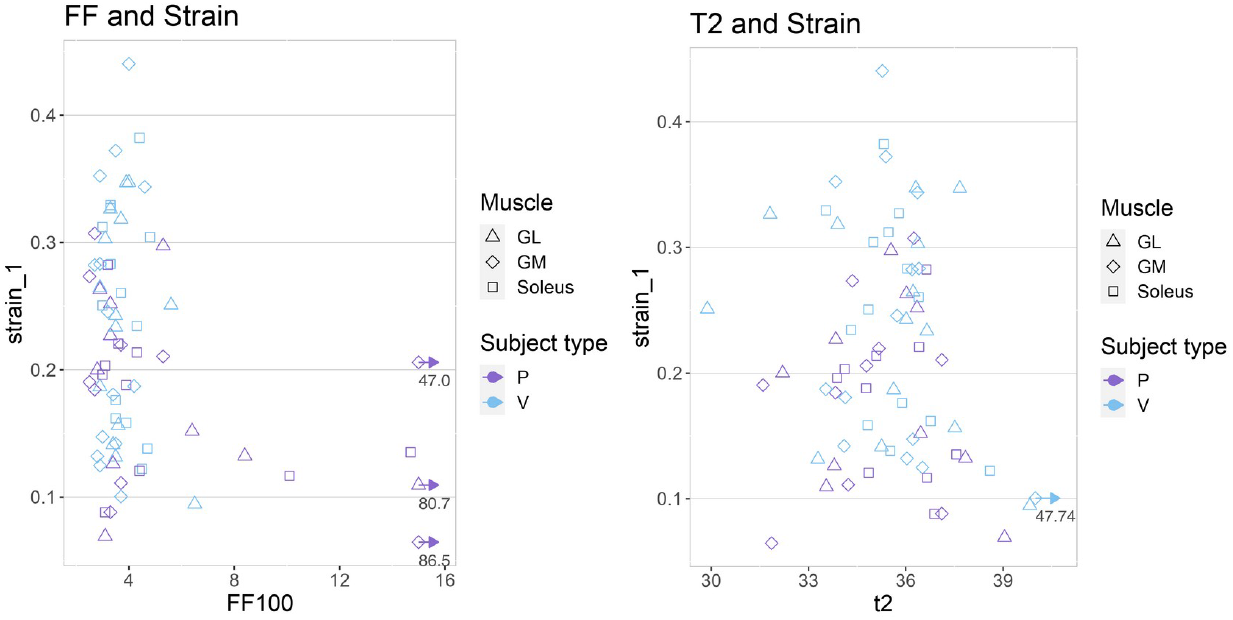
Plots of the strain vs the fat fraction (left) and water T2 (right), for all subject types and muscle ROIs. The correlation between the variables is negligible. Outliers outside the limits of the plot are represented by an arrow and corresponding values.

## Discussion

In this exploratory study, we investigated the potential of dynamic MRI during neuromuscular electrical stimulation to detect differences between HCs and patients affected by metabolic and myotonic myopathies. Previous studies have shown changes in dynamic MRI parameters with age, possibly due to muscle fiber degeneration/loss and other structural abnormalities (19). Since most neuromuscular diseases are associated with muscle fiber loss and fat replacement, and since these structural abnormalities are expected to influence the evaluation of dynamic MRI parameters, we decided to focus on diseases such as myotonias and metabolic myopathies, that are usually not associated with fixed weakness and muscle degeneration, at least in early stages.

Myotonia is characterized, clinically and on neurophysiologic testing, by muscle hyperexcitability and delayed relaxation leading to stiffness or transient weakness, induced by voluntary movement, percussion, changes in temperature, or electrical stimulation. In chloride and sodium channel myotonia, as well as in dystrophic myotonia, the stiffness is prominent with the first movements following a period of rest, and then diminishes and may even disappear. On the contrary, in paramyotonia, stiffness increases with repeated exercise. Beyond stiffness, myotonia may be associated with transient weakness, especially with repeated exercise and especially in the paramyotonia phenotype of sodium channel myotonia. The increase in Na+ currents leading to stiffness or paralysis in sodium channelopathies is more pronounced in type II vs. type I muscle fibers.

The key finding is that strain and strain buildup rate were reduced in the soleus muscle of patients compared to HCs, suggesting that these parameters could serve as biomarkers of muscle dysfunction in muscular and neuromuscular diseases. While other studies detected structural MRI changes in patients with non-dystrophic myotonias (30,31), we did not observe any changes in muscle structure, as assessed by FF/water T2, possibly in view of younger age or milder phenotype, with all but one patient performing well on TUG test. On the contrary, we did not detect changes in FF in patients with myotonia, possibly in view of a milder clinical phenotype. Thus, the lack of significant differences both in conventional imaging and in FF and water T2 measurements between patients and HCs in our study supports the notion that the observed changes truly reflect a change in muscle contractile properties, independently of changes in muscle structure/composition detectable by currently established methods.

This finding is in line with existing literature on the subject of dynamic muscle imaging. Although no comparable study on patients with muscular or neuromuscular diseases has been conducted so far, results from studies in an aging population (16,18,19), and induced atrophy (32) show similar trends in the investigated markers.

The previous study on facioscapulohumeral dystrophy (22) did not show the trend that we could find here when comparing HCs with patients; however, the previous study had the serious limitation of being limited to a single acquired dynamic slice, which reduced its comprehensiveness in evaluating the muscle, and introduced the additional uncertainty of the slice placement in the measurement.

Although all three muscles of the triceps surae show a similar trend, the difference between patients and HCs is more pronounced in the soleus muscle. This can be explained by a placement of the distal electrode just below the belly of the gastrocnemii, thus favoring soleus activation.

This work has several limitations. Most importantly, the group of patients is rather small and heterogeneous, dictated by the availability of the patient pool in the institution where the scans took place. This is a common limitation in the investigation of rare diseases, and future work needs to be done to standardize acquisition and reconstruction pipelines to enable larger, multicentric studies (33). Focusing on larger patient groups with the same pathology will help better define the applicability of dynamic MRI for the evaluation of neuromuscular diseases. Moreover, some of the chosen phenotypes (e.g., myotonia and paramyotonia) are expected to behave in an opposite way after voluntary contraction or repetitive electrical stimulation, with paramyotonia worsening and myotonia improving with repetition. Thus, the assessment of build-up, strain, and release within the same time frame may nullify the differences, if any, meaning that if the changes in contractility are not synchronous, and if the time frame is too long, an average is observed. Another potential source of variability is that the selected disorders could be that myotonias may be associated with increased muscle bulk secondary to hypertrophy from the involuntary exercise caused by myotonic contractions; we do not know the impact of this increased bulk on strength and quality of contraction.

Another limitation was that for this protocol, acquisition of the elicited force was only available before the scan, but not during the scan. So, while the standardization of the force, which is currently the most repeatable method for this kind of acquisition (21), was done outside the scanner room, it could not be repeated at the exact moment of scanning. This possibly introduced an additional factor of intersubject variability in the measurements.

As remarked before (22), the negligible correlations found between strain, fat fraction, and water T2 suggest that these parameters provide complementary information on muscle status. While fat fraction reflects the degree of fatty infiltration and water T2 is sensitive to edema and inflammation, strain and strain rate are markers of the active contraction process. Combining these different imaging biomarkers could provide a more comprehensive evaluation of muscle health in muscular and neuromuscular diseases.

In conclusion, this study provides preliminary evidence that dynamic muscle MRI during neuromuscular electrical stimulation can detect abnormalities in muscle contraction in patients with myotonia and metabolic myopathies, independent of structural MRI abnormalities. The strain and buildup rate in the soleus muscle appear particularly promising as potential biomarkers. However, further research is needed to confirm these findings in larger and more homogeneous patient cohorts, optimize the acquisition protocol, and elucidate the relationship between dynamic MRI parameters and clinical outcomes. By combining dynamic and quantitative MRI techniques, a multi-parametric imaging approach could greatly enhance our understanding and management of neuromuscular disorders.

## Data Availability

Tabular data and analysis code are available on Zenodo at the DOI 10.5281/ZENODO.13169702.
Raw imaging data are available upon reasonale request to the authors.

https://zenodo.org/records/13169702

## Acknowledgment

This work was supported by the Italian Ministry of Health (RC 2022-23).

A Large Language Model (Claude v3, model “Opus”, Anthropic, San Francisco, CA, USA) was used to edit and rephrase parts of the discussion of this paper. The authors take full responsibility for the content.

---

**Supplementary Table 1.** Demographic and disease features of the patients and clinical phenotype.

| Patient_ number | Sex | Age Range | Gene | Mutation | Phenotype (?) | Timed Up and Go | MRI FF/water T2 | myotonia treatment |
| --- | --- | --- | --- | --- | --- | --- | --- | --- |
| P01-2 | M | 66-70 | ZFN9 | CCTG repeat Intron 1 > 75 | dystrophic myotonia type 2 | N | FF |  |
| P02-1 | M | 51-55 | CLCN1 | c.2680C>T (p.R894X) | dominant MC | N | - |  |
| P03-1 | M | 21-25 | CLCN1 | c.2680C>T (p.R894X) | dominant MC | N | - |  |
| P04-1 | F | 51-55 | SCN4A | Val1589Met | sodium channel myotonia | N | - |  |
| P05-1 | M | 46-50 | CLCN1 | c.180+3A>T intron 1; 1182_1186delTG GAA, exon 11 | recessive MC | N | - | mexiletine |
| P08-2 | F | 46-50 | ZFN9 | CCTG repeat Intron 1 > 75 | dystrophic myotonia type 2 | N | - |  |
| P10-1 | M | 56-60 | SCN4A | P1313M | PMC | P | FF |  |
| P11-2 | F | 66-70 | DMPK | E1 | dystrophic myotonia type 1 | N | FF |  |
| P12-3 | F | 46-50 | PYGM | R50X / IVS20-1 | McArdle | N | - |  |
| P14-3 | M | 26-30 | PYGM | c.148C>T (p-Arg50X) and c.2262delA (p.Lys754Asnfs*) | McArdle | N | - |  |

Demographic and disease features of the patients and genetic diagnosis. The functional impact of myotonia /myopathy was assessed at the lower limbs only, by the Timed -Up-and-Go test, that was divided as Normal vs Pathologic

## References

1. Blat Y, Blat S. Drug Discovery of Therapies for Duchenne Muscular Dystrophy. J. Biomol. Screen. 2015;20:1189–1203 doi: 10.1177/1087057115586535.

2. Bushby K, Finkel R, Wong B, et al. Ataluren treatment of patients with nonsense mutation dystrophinopathy. Muscle Nerve 2014;50:477–487 doi: 10.1002/mus.24332.

3. Mercuri E, Muntoni F. Muscular dystrophy: new challenges and review of the current clinical trials. Curr. Opin. Pediatr. 2013;25:701–707 doi: 10.1097/MOP.0b013e328365ace5.

4. Douglas AGL, Wood MJA. Splicing therapy for neuromuscular disease. Mol. Cell. Neurosci. 2013;56:169–185 doi: 10.1016/j.mcn.2013.04.005.

5. Arechavala-Gomeza V, Anthony K, Morgan J, Muntoni F. Antisense oligonucleotide-mediated exon skipping for Duchenne muscular dystrophy: progress and challenges. Curr. Gene Ther. 2012;12:152–160 doi: 10.2174/156652312800840621.

6. Carlier PG, Marty B, Scheidegger O, et al. Skeletal Muscle Quantitative Nuclear Magnetic Resonance Imaging and Spectroscopy as an Outcome Measure for Clinical Trials. J Neuromuscul Dis 2016;3:1–28.

7. Tasca G, Pescatori M, Monforte M, et al. Different molecular signatures in magnetic resonance imaging-staged facioscapulohumeral muscular dystrophy muscles. PloS One 2012;7:e38779 doi: 10.1371/journal.pone.0038779.

8. Monforte M, Laschena F, Ottaviani P, et al. Tracking muscle wasting and disease activity in facioscapulohumeral muscular dystrophy by qualitative longitudinal imaging. J. Cachexia Sarcopenia Muscle 2019;10:1258–1265 doi: 10.1002/jcsm.12473.

9. Tasca G, Monforte M, Ottaviani P, et al. Magnetic resonance imaging in a large cohort of facioscapulohumeral muscular dystrophy patients: Pattern refinement and implications for clinical trials. Ann. Neurol. 2016;79:854–864 doi: 10.1002/ana.24640.

10. Mercuri E, Pichiecchio A, Counsell S, et al. A short protocol for muscle MRI in children with muscular dystrophies. Eur. J. Paediatr. Neurol. EJPN Off. J. Eur. Paediatr. Neurol. Soc. 2002;6:305–307 doi: 10.1016/s1090-3798(02)90617-3.

11. Otto LAM, Pol W-L van der, Schlaffke L, et al. Quantitative MRI of skeletal muscle in a cross-sectional cohort of patients with spinal muscular atrophy types 2 and 3. NMR Biomed. 2020;33:e4357.doi: 10.1002/nbm.4357.

12. Santini F, Deligianni X, Paoletti M, et al. Fast Open-Source Toolkit for Water T2 Mapping in the Presence of Fat From Multi-Echo Spin-Echo Acquisitions for Muscle MRI. Front. Neurol. 2021;12 doi: 10.3389/fneur.2021.630387.

13. Marty B, Baudin P-Y, Reyngoudt H, et al. Simultaneous muscle water T2 and fat fraction mapping using transverse relaxometry with stimulated echo compensation. NMR Biomed. 2016;29:431–443.

14. Wang LH, Shaw DWW, Faino A, et al. Longitudinal study of MRI and functional outcome measures in facioscapulohumeral muscular dystrophy. BMC Musculoskelet. Disord. 2021;22:262 doi: 10.1186/s12891-021-04134-7.

15. Muntoni F, Torelli S, Ferlini A. Dystrophin and mutations: one gene, several proteins, multiple phenotypes. Lancet Neurol. 2003;2:731–740 doi: 10.1016/S1474-4422(03)00585-4.

16. Sinha U, Malis V, Csapo R, Narici M, Sinha S. Shear strain rate from phase contrast velocity encoded MRI: Application to study effects of aging in the medial gastrocnemius muscle. J. Magn. Reson. Imaging JMRI 2018;48:1351–1357 doi: 10.1002/jmri.26030.

17. Malis V, Sinha U, Sinha S. 3D Muscle Deformation Mapping at Submaximal Isometric Contractions: Applications to Aging Muscle. Front. Physiol. 2020;11:600590 doi: 10.3389/fphys.2020.600590.

18. Sinha U, Malis V, Csapo R, Moghadasi A, Kinugasa R, Sinha S. Age-related differences in strain rate tensor of the medial gastrocnemius muscle during passive plantarflexion and active isometric contraction using velocity encoded MR imaging: Potential index of lateral force transmission: Strain Rate Mapping of Calf Muscle Fibers. Magn. Reson. Med. 2015;73:1852–1863 doi: 10.1002/mrm.25312.

19. Deligianni X, Klenk C, Place N, et al. Dynamic MR imaging of the skeletal muscle in young and senior volunteers during synchronized minimal neuromuscular electrical stimulation. Magn. Reson. Mater. Phys. Biol. Med. 2020;33:393–400 doi: 10.1007/s10334-019-00787-7.

20. Deligianni X, Pansini M, Garcia M, et al. Synchronous MRI of muscle motion induced by electrical stimulation. Magn. Reson. Med. 2017;77:664–672 doi: 10.1002/mrm.26154.

21. Deligianni X, Hirschmann A, Place N, Bieri O, Santini F. Dynamic MRI of plantar flexion: A comprehensive repeatability study of electrical stimulation-gated muscle contraction standardized on evoked force. PLOS ONE 2020;15:e0241832 doi: 10.1371/journal.pone.0241832.

22. Deligianni X, Santini F, Paoletti M, et al. Dynamic magnetic resonance imaging of muscle contraction in facioscapulohumeral muscular dystrophy. Sci. Rep. 2022;12:7250 doi: 10.1038/s41598-022-11147-2.

23. Santini F, Bieri O, Deligianni X. OpenForce MR: A low-cost open-source MR-compatible force sensor. Concepts Magn. Reson. Part B Magn. Reson. Eng. 2018;48B:e21404 doi: 10.1002/cmr.b.21404.

24. Liang D, King K, Liu B, Ying L. Accelerating SENSE using distributed compressed sensing. In: Proceedings 17th Scientific Meeting, International Society for Magnetic Resonance in Medicine, Honolulu.; 2009. p. 377.

25. Liu J, Rapin J, Chang T, et al. Dynamic cardiac MRI reconstruction with weighted redundant Haar wavelets (abstr). Proc Int Soc Magn Reson Med 2012;67.

26. Welch EB, Smith DS, Avison MJ, Berglund J, Kullberg J, Ahlström H. Fattyriot - Final Winning Entry Of The 2012 Ismrm Challenge On Water-Fat Reconstruction. 2015 doi: 10.5281/ZENODO.16741.

27. Santini F, Wasserthal J, Agosti A, et al. Deep Anatomical Federated Network (Dafne): an open client/server framework for the continuous collaborative improvement of deep-learning-based medical image segmentation. 2023 doi: 10.48550/arXiv.2302.06352.

28. R Development Core Team. R: A Language and Environment for Statistical Computing. Vienna, Austria: R Foundation for Statistical Computing; 2008.

29. Santini F. Code, data and figures for the paper “Dynamic MR of muscle contraction during electrical muscle stimulation as a potential diagnostic tool for neuromuscular disease.” 2024 doi: 10.5281/ZENODO.13169702.

30. Pedersen JJ, Stemmerik MG, Jacobsen LN, et al. Muscle fat replacement and contractility in patients with skeletal muscle sodium channel disorders. Sci. Rep. 2023;13:2538 doi: 10.1038/s41598-023-29759-7.

31. Morrow B, Zampoli M, van Aswegen H, Argent A. Mechanical insufflation-exsufflation for people with neuromuscular disorders. Cochrane Database Syst. Rev. 2013:CD010044 doi: 10.1002/14651858.CD010044.pub2.

32. Malis V, Sinha U, Csapo R, Narici M, Sinha S. Relationship of changes in strain rate indices estimated from velocity-encoded MR imaging to loss of muscle force following disuse atrophy. Magn. Reson. Med. 79:912–922 doi: 10.1002/mrm.26759.

33. Monforte M, Attarian S, Vissing J, et al. 265th ENMC International Workshop: Muscle imaging in Facioscapulohumeral Muscular Dystrophy (FSHD): relevance for clinical trials. 22–24 April 2022, Hoofddorp, The Netherlands. Neuromuscul. Disord. 2023;33:65–75 doi: 10.1016/j.nmd.2022.10.005.

